# Listening to older voices: Results of a cross-sectional survey of older patient-reported experiences of facility-based healthcare in Nouna, Burkina Faso

**DOI:** 10.1101/2021.12.14.21266715

**Authors:** Ellen M Goldberg, Mamadou Bountogo, Guy Harling, Till Barnighausen, Justine I Davies, Lisa R Hirschhorn

**Author notes:** (co-senior).

## Abstract

**Background:** Ensuring responsive healthcare which meets patient expectations and generates trust is important to increase rates of access and retention. This need is important for aging populations where non-communicable diseases (NCDs) are a growing cause of morbidity and mortality.

**Methods:** We performed a cross-sectional household survey including socio-demographic, morbidities, and health system utilization, responsiveness, and quality outcomes in individuals over age 40 in northwestern Burkina Faso. We describe results and use exploratory factor analysis to derive a contextually appropriate grouping of health system responsiveness (HSR) variables. We used linear or logistic regression to explore associations between socio-demographics, morbidities, and the grouped-variable, then between these variables and health system quality outcomes.

**Results:** Of 2,639 eligible respondents, 26.8% had least one NCD, 56.3% were frail or pre-frail and 23.9% had a recent visit, including only 1/3 of those with an NCD. Highest ratings of care experience (excellent/very good) included ease of following instructions (86.1%) and trust in provider skills (81.1%). The HSR grouping with the greatest factor loading included involvement in decision-making, clarity in communication, trust in the provider, and confidence in provider skills, termed as Shared Understanding and Decision Making (SUDM). In multivariable analysis, higher quality of life (OR 1.02,95%CI 1.01-1.04), frailty (OR 1.47,95%CI 1.00-2.16), and SUDM (OR 1.06,95%CI 1.05-1.09) were associated with greater health system trust and confidence. SUDM was associated with overall positive assessment of the healthcare system (OR 1.02,95%CI 1.01-1.03) and met healthcare needs (OR 1.09,95%CI 1.08-1.11). Younger age and highest wealth quintile were also associated with higher met needs.

**Conclusions:** Recent healthcare access was low for people with existing NCDs, and SUDM was the most consistent factor associated with higher health system quality outcomes. Results highlight the need to increase continuity of care for aging populations with NCDs and explore strengthening SUDM to achieve this goal.

**What is already known?:**

- Health system responsiveness and health system quality outcomes including meeting health needs and trust in the system are important to ensure patient centered care and increase access and retention.
- The process and outcomes of care experience of older adults in Burkina Faso and factors associated with ratings has not been widely studied, information needed to inform efforts to improve engagement in care particularly for individuals with non-communicable chronic diseases (NCDs)
- Shared decision making is emerging as an important component of care to improve engagement in care for people with chronic diseases with studies largely from high income countries

**What are the new findings?:**

- We describe the patterns and gaps in care seeking of people age 40 or older in Burkina Faso, particularly those with non-communicable chronic diseases.
- Highest ratings of care experience included ease of following instructions and trust in provider skills, with lowest ratings in clarity of communication and involvement in decision making.
- We identified a grouped variable was identified using exploratory factors analysis, shared understanding and decision making (SUDM), which was associated with overall positive assessment of the healthcare system and met healthcare needs.

**What do the new findings imply?:**

- Work is needed to increase longitudinal engagement in care for older adults, particularly those with NCDs,
- SUDM may offer an area for strengthening patient-centeredness of care to achieve these goals, but further research is needed to understand the relationships between SUDM and care outcomes, and the impact of strengthening in Burkina Faso.

## Introduction

As access to care has improved in low and middle income country (LMIC) settings, understanding and ensuring the quality of this care has emerged as a critical step to reach effective universal health coverage and health-related sustainable development goals.(1) The Institute of Medicine has defined six domains of quality, including effectiveness (often measured by technical quality), safety, timeliness, equity, efficiency and patient-centeredness.(2) Patient-centeredness has been further emphasized through the World Health Organization’s (WHO) initiative for Integrated People Centered Health Care, which puts the patient at the center of the health care system.(3)

Poor quality is now a leading cause of preventable mortality, overtaking access as a major cause; poor quality contributes to a persistent equity gap, and results in costs to the individual, health care system and individual society.(1,4) Gaps in quality are particularly apparent in non-communicable diseases (NCDs), which represent a growing burden across all countries as populations age.(5) Multiple studies are now showing the magnitude of gaps in quality of care and resulting clinical outcomes (having a condition recognized and adequately managed) in NCDs and among older individuals (1,6–10)

Receipt of person-centered care has been associated with improved healthcare utilization and better health outcomes and patient safety, while poor experiences and perceived quality due to non-responsive care is associated with delay in accessing or returning to care or bypassing the formal care system, whether because of personal experience or through word-of-mouth. (11–13) Confidence and trust in the health system is also an important outcome of the care system, critical for ensuring willingness to access and return to care, and therefore for the management of chronic conditions which are more frequent in the older populations.(14–16)

Measurement around patient-centeredness builds on the WHO Health Systems Responsiveness Framework which identified seven components of responsive outpatient care: dignity, confidentiality, involvement in decision making (autonomy), communication, choice of provider, prompt attention, and quality of basic amenities.(17) Larson directly linked health system responsiveness to experiential quality and proposed two areas for measurement: (1) patient experience of care, a process measure; and (2) patient satisfaction, a health system quality outcome measure of how well provided care meets patient needs and expectations.(18). The relationship between components of responsiveness of care and the health system quality outcomes is not well described, although recent work from Ghana found that higher reported responsiveness was associated with measures of outcomes including reported met medical needs (a measure of satisfaction) and confidence in the health care system.(19)

As health burden and care needs continue to shift to older individuals and those with NCDs, there is a need to expand the measurement of quality beyond providing technically correct care to care which is also empowering and meets older patients needs. (20) Including older individuals in decisions around their care through shared decision making (SDM) is particularly important, not least because it appears to be important for improving self-management and care outcomes.(21) SDM involves the patient and provider collaborating through better communication to identify preferences and making treatment choices that meet the patient’s goals. This approach addresses health system responsiveness domains including autonomy, communication and trust between the patient and provider.

Burkina Faso is one of the poorest countries in the world, with 43.8% of the population live in extreme poverty. (22) Health care expenditure as a percentage of GDP has increased since 2000, reaching 7% by 2016, but out of pocket sources still contribute a large amount to healthcare funding.(23) Although NCDs are estimated to account for up to one-third of deaths in Burkina Faso (24), health services have historically been tailored towards maternal and child health and infectious diseases. Nevertheless, there is increasing attention given to NCDs, including establishment of an NCD division in Ministry of Health (MOH) and a national integrated NCD policy.(25)

We describe the causes of recent healthcare seeking and reported experiences of care in public sector primary and secondary level facilities among older adults in Nouna, a rural region in Burkina Faso. These results are important for providers and policy makers in Burkina Faso and similar settings to facilitate improved experiences of care to increase care seeking and retention of the aging population and begin to reverse the growing burden of NCD-related morbidity and mortality NCDs in Nouna and similar settings in the region.

## Methods

### Study setting

The study was set in the Nouna Health and Demographic Surveillance System (HDSS) area, led by the Centre de Recherche en Santé de Nouna (CRSN) in the Boucle du Mouhoun region, north-western Burkina Faso. The demographic surveillance area of the Nouna HDSS consists of the market town of Nouna and 59 surrounding villages with a total population of 107 000.(26)The formal public health system within the district level includes primary care centers (known as a Center for Health and Social Promotion (CSPS) and a district hospital (known as a medical center with surgical antennae) as well as private clinics and pharmacies.

### Data collection

Data for this cross-sectional study were obtained during the baseline wave of the CRSN Heidelberg Aging Study (CHAS) and has been described in detail elsewhere.(27). Briefly, we randomly sampled 4000 older adults (over 40 years old) from the 2015 CRSN census population. In villages with more than 90 adults over the age of 40, a random sample of households with at least one age-eligible person was created, and one age-eligible adult in each selected household was randomly selected to complete the survey. In villages with fewer than 90 adults over the age of 40, all households with one or more age-eligible individual were included. Data were collected using Open Data Kit (ODK) software on tablet computers at the participants’ houses between May and July 2018. Interviews were conducted either in French or translated into the local languages of Dioula or Mooré by the interviewers.

The household survey contained questions on sociodemographics; self-reported presence of diseases or other health conditions; facility-type last attended; reasons for last health facility visit; reasons for not attending a facility in the last three months; and selected measures of health system responsiveness and health system quality outcomes (Table 1). Other measures included Anxiety (measured using the Generalized Anxiety Disorder question (GAD-2) score),(28) depression (using Patient Health Questionnaire (PHQ-9))(29) and Quality of life (measured using the validated EuroHIS 8-item version of WHOQOL)(30). Disability was measured using the 12 item WHO Disability Assessment Schedule, version 2 (WHODAS-II) disability score,(31). Cognitive functioning was assessed using CSI-D.(32) The Fried frailty score was constructed as described previously.(33)

**Table 1:** sociodemographics, health conditions, reported medical care seeing and health system quality outcomes among individuals who attended versus did not attend a public primary or secondary level facility in the last visit 3 months prior to the survey

|  |  | Overall population | Attended facility in last 3 months | Did not attend facility in last 3 months | P-value |
| --- | --- | --- | --- | --- | --- |
|  | Group | N (%) | N (%) | N (%) |  |
| Total |  | 2639 | 632 | 2007 |  |
| Sex | Female | 1315 (49.8) | 338 (53.5) | 977 (48.7) | 0.035 |
| Age | 40-49 | 1141 (43.2) | 271 (42.9) | 870 (43.3) | 0.079 |
|  | 50-59 | 755 (28.6) | 148 (23.4) | 607 (30.2) |  |
|  | 60-69 | 475 (18) | 145 (22.9) | 330 (16.4) |  |
|  | 70-79 | 217 (8.2) | 61 (9.7) | 156 (7.8) |  |
|  | 80+ | 53 (2) | 11 (1.7) | 42 (2.1) |  |
| Educational attainment | No formal schooling | 2215 (83.9) | 515 (81.5) | 1700 (84.7) | 0.055 |
|  | Some education | 424 (16.1) | 117 (18.5) | 307 (15.3) |  |
| Marital status | Widowed/divorced/single | 606 (23) | 164 (25.9) | 442 (22) | 0.041 |
|  | Married or cohabitating | 2033 (77) | 468 (74.1) | 1565 (78) |  |
| Wealth quintile | 1 (least wealthy) | 499 (18.9) | 103 (16.3) | 396 (19.7) | <0.0001* |
|  | 2 | 522 (19.8) | 103 (16.3) | 419 (20.9) |  |
|  | 3 | 525 (19.9) | 124 (19.6) | 401 (20) |  |
|  | 4 | 549 (20.8) | 132 (20.9) | 417 (20.8) |  |
|  | 5 (most wealthy) | 544 (20.6) | 170 (26.9) | 374 (18.6) |  |
| Self-reported non-communicable diseases (NCD)<sup>†</sup> | Hypertension | 463 (17.5) | 171 (27.1) | 292 (14.5) |  |
|  | Diabetes | 62 (2.3) | 18 (2.8) | 44 (2.2) |  |
|  | Hypercholesterolemia | 11 (.4) | 7 (1.1) | 4 (.2) |  |
|  | Heart disease | 163 (6.2) | 61 (9.7) | 102 (5.1) |  |
|  | Stroke | 36 (1.4) | 12 (1.9) | 24 (1.2) |  |
|  | Chronic respiratory disease | 92 (3.5) | 33 (5.2) | 59 (2.9) |  |
|  | Cancer | 14 (.5) | 9 (1.4) | 5 (.2) |  |
|  | ≥1 NCD | 708 (26.8) | 250 (39.6) | 458 (22.8) | <0.0001* |
| Self-reported TB or HIV<sup>†</sup> | TB | 26 (1) | 12 (1.9) | 14 (.7) |  |
|  | HIV | 16 (.61) | 6 (.9) | 10 (.5) |  |
|  | HIV and/or TB | 41 (1.55) | 17 (2.69) | 24 (1.2) | 0.0001* |
| Self-reported other conditions for > 3 months<sup>†</sup> | Cough | 17 (.6) | 8 (1.3) | 9 (.4) |  |
|  | Headache or dizziness | 50 (1.9) | 16 (2.5) | 34 (1.7) |  |
|  | Musculoskeletal or back pain | 189 (7.2) | 60 (9.5) | 129 (6.4) |  |
|  | Dental | 17 (.6) | 5 (.8) | 12 (.6) |  |
|  | Gastrointestinal | 85 (3.2) | 40 (6.3) | 45 (2.2) |  |
|  | ≥1 other condition | 502 (19) | 181 (28.6) | 321 (16) | <0.0001* |
| Symptoms of mental health disorders (MHD)† | Cognitive impairment on testing | 163 (6.2) | 36 (5.7) | 127 (6.3) |  |
|  | Symptoms of anxiety on testing | 301 (11.4) | 89 (14.1) | 212 (10.6) |  |
|  | Depressive symptoms on testing | 202 (7.7) | 55 (8.7) | 147 (7.3) |  |
|  | ≥1 MHD | 518 (19.6) | 142 (22.5) | 376 (18.7) | 0.039 |
| Frailty | Not frail | 1163 (44.1) | 233 (36.9) | 930 (46.3) | < 0.0001* |
|  | Frail/pre-frail | 1476 (55.9) | 399 (63.1) | 1077 (53.7) |  |
| Disability | WHO DAS s†† | 8.3 (0 - 20.8) | 14.6 (4.2 - 27.1) | 6.3 (0 - 18.8) | <0.0001* |
| Quality of life | WHO QoL †† | 59.4 (46.9 - 65.6) | 56.3 (43.8 - 65.6) | 59.4 (46.9 - 68.8) | <0.0001* |
| Facility type for last visit | Center for Health and Social Promotion | 2206 (83.6) | 496 (78.5) | 1710 (85.2) | <0.0001* |
|  | District Hospital | 433 (16.4) | 136 (21.5) | 297 (14.8) |  |
| Financial access | Did not borrow or sell anything | 2250 (85.3) | 539 (85.3) | 1711 (85.3) | 0.98 |
|  | Borrowed or sold something to attend clinic | 389 (14.7) | 93 (14.7) | 296 (14.7) |  |
| Outcomes |  |  |  |  |  |
| Reported met need | Excellent | 234 (8.9) | 56 (8.9) | 178 (8.9) | 0.0058* |
|  | Very Good | 968 (36.7) | 262 (41.5) | 706 (35.2) |  |
|  | Good | 1293 (49) | 275 (43.5) | 1018 (50.7) |  |
|  | Fair | 116 (4.4) | 33 (5.2) | 83 (4.1) |  |
|  | Poor | 28 (1.1) | 6 (.9) | 22 (1.1) |  |
| Trust and confidence in health care system | Very confident | 872 (33) | 246 (38.9) | 626 (31.2) | 0.0003* |
|  | Somewhat confident | 1610 (61) | 358 (56.6) | 1252 (62.4) |  |
|  | Not very confident | 138 (5.2) | 26 (4.1) | 112 (5.6) |  |
|  | Not at all confident | 19 (.7) | 2 (.3) | 17 (.8) |  |
| Overall view of the health care system in this country | Positive | 1612 (61.1) | 408 (64.6) | 1204 (60) |  |
|  | Neutral | 956 (36.2) | 212 (33.5) | 744 (37.1) |  |
|  | Poor | 71 (2.7) | 12 (1.9) | 59 (2.9) | 0.040 |
| † P value represents ≥1 condition versus none or HIV/TB using chi square |  |  |  |  |  |
| †† Scale from 0-100, median (IQR) |  |  |  |  |  |
| * P < 0.05 when adjusted for multiple comparisons using a Bonferroni correction |  |  |  |  |  |
| Table created by authors |  |  |  |  |  |

### Definition of variables

#### Health System Responsiveness and Health System quality outcomes

A subset of all possible health system responsiveness domains was included due to constraints of the survey length. Questions were selected based on discussion between investigators and their perceived relevance to the local context and focus on experiential quality. They were taken from published studies in sub-Saharan Africa (Appendix Table 1A).(Baltussen, 2002; Miller et al., 2014; Ratcliffe et al., 2020) and only asked for individuals with recent (within last three months) visit. Health system quality outcome questions included trust and confidence in receiving effective treatment, patient satisfaction (how well the received care met health need), and the overall view of the health system.

#### Demographic characteristics

Marital status was categorized as married/cohabiting versus single/widowed/divorced. Educational level was dichotomized as no education or any education. Participants were asked 37 questions on household assets and dwelling characteristics; from these, wealth quintiles were derived from the Filmer and Pritchett first principal component method.(34) Age was categorized in 10-year groups for the descriptive and univariate analysis and as a continuous variable in the multivariable analysis.

#### Disease categories

We included several self-reported conditions including non-communicable conditions (hypertension, diabetes, hypercholesteremia, heart disease, stroke, chronic respiratory disease, and epilepsy), and communicable diseases (HIV and tuberculosis (TB)). Self-reported chronic symptoms (lasting for more than 3 months) included cough, headache, musculoskeletal or back pain, dental, or gastrointestinal manifestations. Some health conditions were captured as free text; these were translated and categorized through discussions among authors where necessary.

Participants were defined as having symptoms of anxiety based on a GAD-2 score ≥3, depression based on PHQ-9 score ≥ 10 and cognitive functioning as possible/probable cognitive impairment for CSI-D score <7. Participants with at least one symptom of anxiety, depression, or cognitive impairment on testing were defined as having a neurological or mental health diagnosis. WHODAS-II and quality of life were normalized to 0-100. For frailty, participants were dichotomized as robust versus prefrail/frail/unable to complete assessment.

## Analysis

### Analytic sample

We limited our sample to those who sought care at their last visit from a public sector primary (Center for Health and Social Promotion or secondary level (District Hospital) facility, to reflect our focus on local care seeking and the most common sources of care (93% of individuals for the variables of interest (see CONSORT diagram Appendix Figure 1A). Using unweighted data, we described demographic characteristics, disease state, visit characteristics, and health system outcomes both among the whole sample surveyed and separately for participants who recently sought care (within the last 3 months) and those who did not. We then determined bivariate associations between these characteristics and health system quality outcomes for those who sought care versus those who did not using chi-squared, ranksum, or t-test as appropriate. We used a Bonferroni correction to adjust for multiple comparisons.

### Health System Responsiveness and Health System Quality Outcomes

We conducted an exploratory factor analysis of the experiential quality questions (Appendix Figure 2A) to explore grouping of these variables, based on our assumption that one or more common constructs related to engagement in care and health system quality outcomes underlay our observed variables. We then used the variable with the greatest factor loading (“the HSR-group variable”) in subsequent analyses by scaling each individual component to 0-100 with 0 representing the lowest and 100 the highest rating and averaged them to arrive at a final variable between 0-100.

### Bivariate analyses

We described individual responsiveness ratings among recent care seekers. We limited these analyses to individuals with a visit in the last 3 months to reduce recall bias. We then tested for bivariate associations between demographic characteristics, health status (one or more self-reported NCD, one or more self-reported “other” condition, one or more symptom of mental health disorder, quality of life, frailty and disability) facility type, financial access, wait time, and the HSR-group variable and the three health system quality outcomes. We did a similar analysis with the HSR-group variable as the outcome of interest.

### Multivariable analyses

We ran logistic regression and linear regression for health system quality outcomes and the HSR-group variable respectively. Variables that met an inclusion criterion of P < 0.2 in the bivariate analyses were included. We also included age, sex, educational attainment, and wealth quintile, given their associations with reported experiential quality and selected health systems quality outcomes in previous studies.(19,35–37).

All statistical analyses were performed using Stata software (version 15.1; StataCorp LLC, College Station, Texas).

IRB: Ethical approval was obtained from Ethics Commission of the medical faculty Heidelberg (S-120/2018), the Burkina Faso Comité d’Ethique pour la Recherche en Santé (CERS) in Ouagadougou (2018-4-045) and the Institutional. Ethics Committee (CIE) of the CRSN (2018-04). Oral assent was sought from all village elders. Written informed consent was obtained from each participant and a literate witness assisted in cases of illiteracy.

Patients or the public were not involved in the design, or conduct, or reporting, or dissemination plans of our research

## Results

### Population

Overall, 3,028 individuals responded to the survey including questions about care seeking with 177 excluded for missing data and 212 for care at a private sector facility or tertiary care hospital (Appendix Figure 1). Among the 2,639 who reported their last visit to a public sector primary or secondary level facility, 632 (23.9%) sought care at one of these facilities in the 3 months prior to the survey (Table 1). Overall, one half (50%) were women, with 42.8% age 40-49 and 10.5% age 70 or older. Education was low (83.8% reported no formal education), and three quarters (76.4%) were married or cohabitating. One quarter reported at least one NCD (26.8%), with lower rates of communicable diseases such as HIV or TB (2.8%). The median WHO DAS score was 8.3 (interquartile range (IQR) 0-22.9) and for QoL was 59.4 (IQR 46.9-65.6), while 56.3% were categorized as frail or pre-frail.

Individuals who attended care in the last 3 months were significantly wealthier than those who did not attend care in this timeframe, and more likely to have at least one NCD, have either HIV or TB or both, or other conditions lasting for more than 3 months Despite individuals with chronic diseases having attended clinic more recently, 65% of respondents with these conditions did not report attending care in this timeframe, including 62.7% of patients reporting hypertension and 66.7% of individuals reporting diabetes.

People who had attended in the last 3 months also had significantly higher disability measured by DAS scores (14.6 versus 6.3), lower QoL (56.3 versus 59.4), and were more likely to be frail (63.1% versus 53.7%) than those with no visits in the last 3 months; all p<.0001.

#### Visits characteristics

The most common reasons overall for seeking care were for acute conditions (79.1%) including fever or malaria (51.6%), musculoskeletal pain (9.6%), and diarrhea or stomach-ache (8.4%). Chronic conditions accounted for care seeking in 12.9% including hypertension (6.2%), other cardiac conditions (2.1%) and diabetes (0.6%) (Appendix Table 2A) The most common reasons for care-seeking within the past 3 months were fever or malaria (37.8), high blood pressure (12.8%), musculoskeletal pain (12.0%), complaints related to the ear, nose or throat (7.4%), or diarrhea or stomach-ache (7.0%). Not being sick was the most frequent reason for no recent care-seeking (87.3%) (Appendix Table 3A). Among those who stated other reasons for not seeking care, cost was the most common reason (50.4%), followed by preferring to see a traditional healer (11.6%) and poor previous experiences with the health system (6.0%).

**Table 2:** Experience at last visit to a public sector primary or secondary level facility in the 3 months prior to survey.

| Measure | Rating | N (%) |
| --- | --- | --- |
| Clarity | Excellent | 45 (7.1) |
|  | Very Good | 260 (41.1) |
|  | Good | 287 (45.4) |
|  | Fair | 39 (6.2) |
|  | Poor | 1 (.2) |
| Ease of following | Excellent | 81 (12.8) |
|  | Very Good | 463 (73.3) |
|  | Good | 76 (12) |
|  | Fair | 11 (1.7) |
|  | Poor | 1 (.2) |
| Provider medical knowledge and skills | Excellent | 64 (10.1) |
|  | Very Good | 259 (41) |
|  | Good | 280 (44.3) |
|  | Fair | 27 (4.3) |
|  | Poor | 2 (.3) |
| Trust in skills and abilities of health workers at the facility | Very much | 104 (16.5) |
|  | Quite a bit | 408 (64.6) |
|  | Some | 105 (16.6) |
|  | Very little | 13 (2.1) |
|  | Not at all | 2 (.3) |
| Involvement in decision making | Excellent | 35 (5.5) |
|  | Very Good | 159 (25.2) |
|  | Good | 268 (42.4) |
|  | Fair | 101 (16) |
|  | Poor | 69 (10.9) |
| Shared Understanding and Decision Making (SUDM) | Median (Interquartile range) | 62.5 (50 - 75) |
| Borrowed money or sold anything to pay for health care | Yes | 93 (14.7) |
|  | No | 539 (85.3) |
| Wait time (median, IQR) |  | 20 (10 - 30) |
| Table created by authors |  |  |

**Table 3:** Bivariate and multivariable analysis of factors associated with higher shared understanding and decision making (SUDM) among individuals with a visit to a primary or secondary level public sector facility in the 3 months prior to the survey.

| Sex |  | Bivariate analysis odds ratio(95% CI) | P value | Multivariable analysis odds ratio(95% CI) | P value |
| --- | --- | --- | --- | --- | --- |
| Sex | Male | Reference |  | Reference |  |
|  | Female | -1.74 (-4.01-0.54) | 0.13 | **-2.43 (-4.76 - -0.11)** | **0.040** |
| Age (per year) |  | 0.12 (0.016 - 0.22) | 0.023 | 0.072 (-0.046 - 0.19) | 0.23 |
| Educational attainment | No formal schooling | Reference |  | Reference |  |
|  | Some education | 0.32 (-2.61 - 3.24) | 0.83 | -0.57 (-3.64 - 2.50) | 0.72 |
| Marital status | Widowed/divorced/single | Reference |  |  |  |
|  | Married/cohabitating | -0.44 (-3.03 - 2.15) | 0.74 |  |  |
| Wealth quintile† | 1 | Reference |  | Reference |  |
|  | 2 | 0.24 (-3.87 - 4.36) | 0.91 | -0.33 (-4.28 - 3.62) | 0.87 |
|  | 3 | 0.37 (-3.45 - 4.19) | 0.85 | 0.15 (-3.64 - 3.94) | 0.94 |
|  | 4 | -0.76 (-4.64 - 3.12) | 0.70 | -1.12 (-4.88 - 2.64) | 0.56 |
|  | 5 | 1.38 (-2.31 - 5.07) | 0.46 | -0.13 (-3.87 - 3.60) | 0.95 |
| Facility type | Center for Health and Social Promotion | Reference |  | Reference |  |
|  | **Medical Center with Surgical Antenna** | **6.04 (3.32 - 8.76)** | **<0.001** | **5.48 (2.58 - 8.38)** | **< 0.001** |
| Financial Accessibility | Borrowed or sold anything to attend clinic | Reference |  |  |  |
|  | Did not borrow or sell anything | 1.06 (-2.14 - 4.27) | 0.56 |  |  |
| Non-communicable diseases (NCD) | No NCDs | Reference |  | Reference |  |
|  | ≥1 NCD | 1.94 (-0.38 - 4.26) | 0.10 | 1.28 (-1.09 - 3.64) | 0.29 |
| TB or HIV | No TB or HIV | Reference |  |  |  |
|  | TB and/or HIV | -4.05 (-11.07 - 2.96) | 0.26 |  |  |
| Other conditions for > 3 months | No other conditions | Reference |  |  |  |
|  | ≥1 other condition | -0.88 (-3.39 - 1.63) | 0.49 |  |  |
| Mental health disorders (MHD) | No MHDs | Reference |  | Reference |  |
|  | ≥1 MHD | 2.51 (-0.20 - 5.23) | 0.070 | 1.53 (-1.39 - 4.45) | 0.30 |
| Frailty | Not frail/pre-frail | Reference |  |  |  |
|  | Frail | 1.16 (-1.19 - 3.51) | 0.33 |  |  |
| Disability | WHO DAS score†† | 0.063 (0.00077 - 0.12) | 0.047 | 0.013 (-0.06 - 0.09) | 0.74 |
| Quality of life | WHO QoL score †† | -0.036 (-0.11 - 0.039) | 0.34 |  |  |
| Table created by authors |  |  |  |  |  |

#### Health System Quality outcomes

Overall, 32.7% of respondents were very confident that if they got sick, the health system could meet their needs. Compared with individuals with a visit over 3 months ago, individuals with recent visits had higher trust and confidence in the health system to provide effective care if they were sick (38.3% versus 30.8% very confident, p<.0004), although rates remained low. No differences were seen in their needs being met from their last visit or in overall opinion of the national health system (Table 1).

#### Experiences of care at facilities (Health System Responsiveness Variables)

Among individuals with a visit to a public sector primary (CSPS) or secondary level (district hospital) public facility in the last 3 months, the median wait time was 20 minutes (IQR 10-30) while time spent with the provider was 15 minutes (IQR 10-25). Financial access was a challenge with 14.7% borrowing money or selling something to pay for health care. The highest ratings of experience of care (defined as excellent or very good) were in ease of following instructions (86.1%) and trust in the skills and abilities of the facility providers (81.1%). Lower ratings were seen for provider medical knowledge and skills (51.2%), clarity of communications (48.2%), with the lowest ratings in involvement in decision making (30.7%) (Table 2).

The variable grouping with the greatest factor loading (the HSR-group variable) combined the results for questions on involvement in decision-making (autonomy), clarity in communication, trust in the provider, and confidence in providers’ skills (factor loadings of 0.44, 0.73, 0.57, and 0.69, respectively) (Appendix). After discussion between authors, we agreed that these variables reflected components necessary for shared understanding and decision making and termed the resultant variable as such (SUDM). We used the scaled variable as described in the methods and chose to not weight variable components as all were assumed to be equally important for SUDM. The median score for SUDM was 58.3 (Interquartile range (IQR) 50 - 75). In a multivariable analysis, only being seen in a district hospital was associated with higher SUDM (Adjusted odds ratio (aOR) 5.91 (95% CI 2.87 - 8.96)) (Table 3).

#### Factors associated with health system quality outcomes

In the multivariable analysis higher quality of life (OR 1.02, 95% CI 1.01-1.04), frailty (OR 1.47, 95% CI 1.00-2.16) and SUDM (OR 1.06, 95% CI 1.05-1.09) were all associated with greater trust and confidence in the health system to provide effective care if they were sick (Table 4). SUDM was associated with overall positive assessment of the health care system in Burkina Faso (OR 1.02, 95% CI 1.01-1.03) and met healthcare needs in the last visit (OR 1.09, 95% CI 1.08-1.11). Younger age and highest wealth quintile were also associated with higher scores for met needs, while having at least one mental health condition was associated with less positive ratings of the overall health system.

**Table 4:** Multivariable regression for health system quality outcomes among respondents with a visit to a public sector primary or secondary level facility in the 3 months prior to the survey

|  |  | Trust and Confidence in healthcare system |  | Overall view of healthcare system |  | Health care needs met |  |
| --- | --- | --- | --- | --- | --- | --- | --- |
|  |  | Bivariate analysis | Multivariable analysis | Bivariate analysis | Multivariable analysis | Bivariate analysis | Multivariable analysis |
|  |  | OR (95% CI) | OR (96% CI) | OR (95% CI) |  | OR (95% CI) |  |
| Sex | Male | Reference | Reference | Reference | Reference | Reference | Reference |
|  | Female | 0.73<br>(0.53 - 1.01) | 0.83<br>(0.58 - 1.19) | 0.89<br>(0.64 - 1.23) | 0.99<br>(0.70 - 1.40) | 0.86<br>(0.63 - 1.17) | 1.03<br>(0.71 - 1.49) |
| Age* |  | 1.00<br>(0.98 - 1.01) | 1.00<br>(0.98 - 1.01) | 1.00<br>(0.99 - 1.01) | 1.01<br>(0.99 - 1.03) | 0.99<br>(0.98 - 1.01) | **0.98**<br>**(0.97 - 1.00)** |
| Education | No formal schooling | Reference | Reference | Reference | Reference | Reference | Reference |
|  | Some education | 1.02<br>(0.68 - 1.54) | 0.97<br>(0.60 - 1.56) | 0.93<br>(0.61 - 1.42) | 1.07<br>(0.67 - 1.70) | 1.09<br>(0.73 - 1.63) | 0.92<br>(0.56 - 1.50) |
| Wealth quintile | 1 | Reference | Reference | Reference | Reference | Reference | Reference |
|  | 2 | 1.58<br>(0.90 - 2.79) | 1.49<br>(0.79 - 2.81) | 1.00<br>(0.56 - 1.77) | 1.00<br>(0.55 - 1.80) | 1.17<br>(0.68 - 2.03) | 1.24<br>(0.65 - 2.35) |
|  | 3 | 1.86<br>(1.08 - 3.21) | 1.60<br>(0.87 - 2.94) | 1.05<br>(0.61 - 1.82) | 1.03<br>(0.58 - 1.82) | 1.53<br>(0.90 - 2.58) | 1.58<br>(0.85 - 2.94) |
|  | 4 | 1.13<br>(0.66 - 1.96) | 0.99<br>(0.53 - 1.83) | 1.24<br>(0.71 - 2.14) | 1.27<br>(0.72 - 2.25) | 1.19<br>(0.71 - 2.00) | 1.25<br>(0.68 - 2.30) |
|  | 5 | 1.31<br>(0.78 - 2.20) | 0.88<br>(0.48 - 1.63) | 0.77<br>(0.46 - 1.27) | 0.77<br>(0.44 - 1.34) | 1.83<br>(1.11 - 2.99) | **1.85**<br>**(1.00 - 3.42)** |
| Facility | CSPS | Reference |  | Reference | Reference | Reference | Reference |
|  | District Hospital | 1.13<br>(0.77 - 1.66) |  | 0.68<br>(0.46 - 1.00) | 0.70<br>(0.46 - 1.08) | 1.43<br>(0.98 - 2.10) | 0.90<br>(0.56 - 1.45) |
| Financial access | Did not borrow/sell anything | Reference |  | Reference |  | Reference |  |
|  | Borrowed/sold something | 0.94 (0.60 - 1.48) |  | 0.85 (0.54 - 1.33) |  | 1.24 (0.80 - 1.92) |  |
| NCDs | None | Reference |  | Reference |  | Reference |  |
|  | ≥1 NCD | 1.05<br>(0.76 - 1.45) |  | 0.86<br>(0.61 - 1.19) |  | 1.21<br>(0.88 - 1.67) |  |
| HIV or TB | None | Reference | Reference | Reference |  | Reference |  |
|  | HIV and/or TB | 0.47<br>(0.15 - 1.47) | 0.64<br>(0.19 - 2.18) | 0.77<br>(0.29 - 2.08) |  | 0.53<br>(0.19 - 1.45) |  |
| MHD | None | Reference |  | Reference | **Reference** | Reference |  |
|  | ≥1 MHD | 1.07<br>(0.73 - 1.56) |  | 0.51 (0.35 - 0.74) | **0.52**<br>**(0.34-0.80)** | 1.18<br>(0.81 - 1.72) |  |
| Frailty | Not frail/pre-frail | Reference | **Reference** | Reference | Reference | Reference |  |
|  | Frail | 1.32<br>(0.95 - 1.85) | **1.47**<br>**(1.00-2.16)** | 0.75<br>(0.53 - 1.06) | 0.83<br>(0.57 - 1.21) | 1.12<br>(0.81 - 1.55) |  |
| Disability (DAS) | DAS score<sup>††</sup> | 1.00<br>(0.99 - 1.01) |  | 0.99<br>(0.98 - 1.00) | 0.99<br>(0.98-1.01) | 1.00<br>(0.99 - 1.01) |  |
| QOL | QoL score<sup>††</sup> | 1.02<br>(1.01 - 1.03) | **1.02**<br>**(1.01 - 1.04)** | 1.00<br>(0.99 - 1.01) |  | 1.01<br>(0.99 - 1.02) |  |
| Wait time |  | 0.64<br>(0.18 - 2.27) |  | 2.18 (0.61 - 7.78) |  | 1.19<br>(0.34 - 4.12) |  |
| SUDM |  | 1.06<br>(1.05 - 1.07) | **1.06**<br>**(1.05 - 1.09)** | 1.02<br>(1.00 - 1.03) | **1.02**<br>**(1.01-1.03)** | 1.09<br>(1.07 - 1.11) | **1.09**<br>**(1.08 - 1.11)** |

\*per year
CSPS: Center for Health and Social Promotion, NCD: Non communicable diseases, MHD: Mental health Disorder, QOL: Quality of Life SUDM: Shared Understanding and Decision making
Table created by authors

## Discussion

Ensuring longitudinal preventive, promotive and curative primary care among older adults in resource constrained settings is critical to reduce the burden of morbidity and mortality related to NCDs. In this household survey of older individuals in Nouna, Burkina Faso, we found that about one-quarter of individuals sought care at a public primary or secondary care facility in the last three months. Individuals who reported higher wealth, presence of an NCD or communicable disease, high disability and frailty and lower quality of life were more likely to have received recent care. SUDM was found to be the most consistent factor associated with higher health system quality outcomes including satisfaction, confidence in the health systems and overall quality of care.

Acute conditions were the most common reason for care seeking among this older population overall, with one-fifth of recent care seeking for more chronic conditions. However, only one third of patients who self-reported an NCD and 41% of those with TB or HIV had a visit in the last three months, despite recommendations from many institutions including the World Health Organization that individuals with NCDs be seen at least every three months.(38) The lack of recent visits for individuals with chronic conditions requiring longitudinal care is of concern given the importance of ongoing management even when symptoms are not present. Data from Serra Leone found that knowledge about cardiovascular disease risk factors and costs were barriers to accessing care(39), but similar insights from Burkina Faso and among older adults were not found. More work to understand the scope and causes of this challenge in similar settings is needed to develop effective interventions to strengthen the quality of primary and secondary care to ensure not just once-off access but continuity and comprehensiveness of care, core dimensions of effective primary care.(40)(41)

Shared decision making is defined as “process jointly shared by patients and their health care provider”. It aims at helping patients play an active role in decisions concerning their health, which is the ultimate goal of patient-centered care” (42). Shared decision making has been studied since the 1990’s and increasingly important as the push for more people-centered primary care has emerged from the World Health Organizations and the Astana Declaration in 2018.(43) The importance of shared decision making and effective communication for management of chronic conditions has been a focus of research in high income countries with lower rates of shared decision making have been found among older individuals and those with poorer health, and associated with lower adherence to care and treatment.(20,21) Achieving shared decision making requires engagement in decision making, effective communication and good provider-patient relationships, factors which were captured in our SUDM measure. Similar to our study, higher rates of shared decision-making rates have been associated with better satisfaction identifying an area for improving quality and outcomes of care for older individuals and people with NCDs.(44)

Rating of care experience variables, again pointed to areas where change is needed. Participants reported high ratings of some other areas of visit experience (ability to follow advice and trust in provider skills), while other areas were lower, with one-half or fewer reporting high provider technical skills, clarity of communication, or involvement in decision making. Compared with other studies, clarity of communications was lower in our study (48% versus 66-100% in Tanzania and close to 60% in Ghana), although variability in populations, survey questions and scoring makes comparisons challenging.(19)(45) In contrast, in Ghana female patients, gave lower ratings for involvement, although the population was younger overall than in our study.(19)

Trust and confidence in the health system was high, but lower among those not seeking recent care, as well reporting of met needs during the most recent care encounter, offering an opportunity for improving perceptions, engagement-in and delivery-of care. This finding is similar to results from a survey in Ghana of women. In a study in Burkina Faso, perceived quality of care was a determinant for retention in care at a site, important for the continuity needed for NCDs and effective primary care more broadly, and identifying an area where improvement is needed.(46)

While geographic access was only rarely given as a reason for no recent care seeking, 14.5% had to borrow or sell something to attend a clinic, representing a significant burden among a population with high poverty. This measure also may underestimate cost burdens such as individuals who had to forgo consumption of other goods or services such as food to access their health care. While the lower wealth among non-users was similar to findings to Dong et al they also found higher rates of financial access as a barrier than in our study.(47)

Our study had some key limitations. First, we were unable to collect all the dimensions of the traditional health systems responsiveness domains - aspects such as respect and confidentiality might have added to our understanding of care experience in this population. The self-reported nature of past condition information may have underestimated actual prevalence due to absent or forgotten diagnoses. We also limited our analyses to individuals visiting a public sector facility providing primary or secondary level care, excluding the small proportion of participants using private or higher-level facilities, to focus on the local care system delivery. However, given the expanding role of the private sector in many countries, future work focusing on these facilities should be planned.

In conclusion, we provide a comprehensive and mixed picture of public-sector health facility care seeking behaviors and user quality experiences among older individuals in rural Burkina Faso. A minority of individual have sought recent care, most frequently for acute conditions, despite a burden of NCDs which need continuity of care. Among those with recent visits, the importance of shared understanding and engagement in decision making was seen across all measured health systems quality outcomes. Situating our findings was limited by the availability of comparable population-representative samples in rural, low-income settings – efforts to measure similar patient experiences should provide substantial benefit. Our findings provide insights into designing health system and care delivery interventions to improve the experience and involvement in care of the growing elderly population in rural LMICs. These interventions are particularly important for those with chronic conditions for whom ongoing care is critical to reduce preventable mortality and mortality.

## Data Availability

All data produced in the present study are available upon reasonable request to the authors

## Acknowledgements

We would like to thank the team who managed and implemented the survey and the community members who shared their experiences with us and Dr Sie who was integral to the design and performance of the survey.

## Contributors

TB, JD, and GH conceived and designed the overall CSRN CHAS Study. MB and GH coordinated baseline data collection and preparation with support from JD, and LRH contributed to the design of the CSRN CHAS household survey. LRH and JD designed the current study. EMG conducted the analysis, and LRH, JD and EMG interpreted the analysis and developed, led the writing and revisions of the manuscript. All authors substantively reviewed manuscript, inputted into revisions and approved the final manuscript.

