## Appendices for "Listening to older voices: Results of a cross-sectional survey of older patient-reported experiences of facility-based healthcare in Nouna, Burkina Faso": Nouna Appendices 10 OCt .pdf

Table 1A: Experience in care and health system quality outcome questions

| Domain | Question | Response options |
|---|---|---|
| Promptness | Wait time | Minutes |
| Communication<br>• *clarity* | How would you rate the provider's availability to explain things in a way that you could understand? | 5 point likert * |
| • *Ability to follow advice* | How easy or difficult was it for you to follow the provider's advice? | |
| Providers Technical skills | What do you think about the care provider's medical knowledge and skills? | 5 point likert * |
| Trust in provider | How much do you trust the skills and abilities of the health workers at this facility? | 5 point likert * |
| Autonomy/shared decision making | How would you rate your experience of being involved in making decisions for your treatment?) | Poor |
| Financial Access | Did you have to borrow money or sell anything to pay for this health care (including transportation, fees, medication) | Yes/No |
| **Outcomes** | | |
| Trust and confidence in health care system | How confident are you that if you became very sick tomorrow, you would be able to receive effective treatment from the health system? | 4 point liverty *** |
| Met need | Overall, thinking about your entire last visit, please rate how well the care you received met your health needs. That is, how much did the visit help solve your health problem or help you feel better? | 5 point likert * |
| Overall view of the health care system in Burkina Faso | Which of the following statements comes closest to expressing your overall view of the health care system in this country?<br>• Our health care system has so much wrong with it that we need to completely rebuild it (coded as poor)<br>• There are some good things in our health care system, but major changes are needed to make it work better (coded as neutral)<br>• On the whole, the system works pretty well and only minor changes are necessary to make it work better (coded as positive) | Choose one |

*Excellent, Very good, good, fair, poor , ** Very much, quite a bit, some, very little, not at all, *** Very confident, somewhat confident, not very confident, not at all confident

Table created by authors

Figure 1A: Consort Diagram

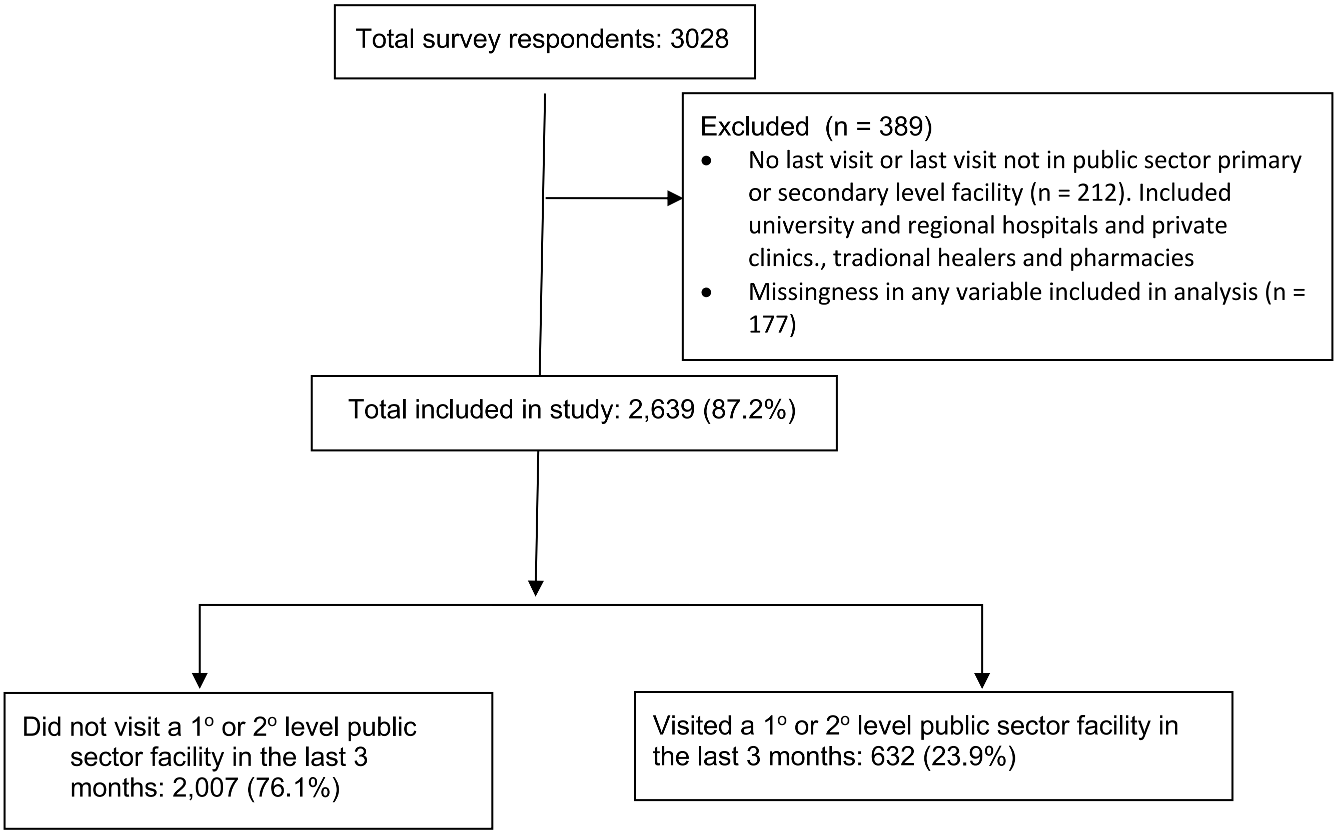

Table 2A: Self reported reasons or last visit among individuas with their last visit at public secto primary or secondary level facility

| | Self-reported Reasons for visit | Total | Faciity visit in last 3 months | Facility visit > 3 months ago |
|---|---|---|---|---|
| | | N (%) | N (%) | N (%) |
| Acute | Fever / Malaria | 1,361 (51.6) | 239 (37.8) | 1,122 (55.9) |
| | Pain in the back, limbs or joints | 254 (9.6) | 76 (12.0) | 178 (8.9) |
| | Diarrhoea / stomach ache | 221 (8.4) | 44 (7.0) | 177 (8.8) |
| | Ear, Nose, Throat | 182 (6.9) | 47 (7.4) | 135 (6.7) |
| | Eye disease | 120 (4.5) | 40 (6.3) | 80 (4.0) |
| | Accident / Injury | 100 (4.5) | 24 (3.8) | 76 (3.8) |
| | Skin problem | 65 (2.5) | 18 (2.8) | 47 (2.3) |
| | Dental problem | 49 (1.9) | 14 (2.2) | 35 (1.7) |
| | Weakness of arm/legs | 25 (0.9) | 12 (1.9) | 13 (0.6) |
| | Respiratory illness | 17 (0.6) | 9 (1.4) | 8 (0.4) |
| | Typhoid fever | 16 (0.6) | 2 (0.3) | 14 (0.7) |
| | Breathlessness | 5 (0.2) | 4 (0.6) | 1 (0.0) |
| | Meningitis | 4 (0.2) | 0 (0) | 4 (0.2) |
| | **Acute (all)*** | **2,087 (79.1)** | **427 (67.6)** | **1,660 (82.7)** |
| Chronic | High blood pressure | 163 (6.2) | 81 (12.8) | 82 (4.1) |
| | Other heart condition | 55 (2.1) | 26 (4.1) | 29 (1.4) |
| | Diabetes, including symptoms | 16 (0.6) | 8 (1.3) | 8 (0.4) |
| | Asthma | 10 (0.4) | 2 (0.3) | 8 (0.4) |
| | Malignant Tumor / Cancer | 7 (0.3) | 4 (0.6) | 3 (0.1) |
| | **Chronic (all)*** | **249 (9.4)** | **119 (18.8)** | **130 (6.5)** |
| Other | Other | 254 (9.6) | 80 (12.7) | 174 (8.7) |
| | Antenatal care, post-natal care, or delivery | 77 (2.9) | 12 (1.9) | 65 (3.2) |
| | Family planning | 17 (0.6) | 4 (0.6) | 13 (0.6) |
| | Peptic ulcer | 36 (1.4) | 18 (2.8) | 18 (0.9) |
| | **Other (all)**** | **382 (14.5)** | **114 (18.0)** | **268 (13.4)** |

*p<.001. **p=.004

Table created by authors

Table 3A: Most common reasons for no visit in the last 3 months

| Most common reasons for no visits (n=232) | N (%) |
|---|---|
| Was not sick | 1,775 (88.4%) |
| Too expensive | 117 (6.0%) |
| Preferred to see a healer | 27 (1.3%) |
| Unskilled staff/do not think staff are good | 2 (0.10%) |
| Ineffective treatment | 12 (0.6%) |
| Bad experience in the past | 11 (0.5%) |
| Preferred to see a community health worker | 10 (0.5%) |
| Long wait | 9 (0.4%) |
| Too far away/could not get transport | 8 (0.4%) |
| Did not have time/childcare | 6 (0.3%) |
| Institution not clean | 1 (0.05%) |
| Other | 59 (2.4%) |

Table created by authors

Figure 2A. Results from Exploratory factor analysis

| Factor | Eigenvalue | Difference | Proportion | Cumulative |
|---|---|---|---|---|
| Factor1 | **1.68102** | **1.18859** | **0.9568** | **0.9568** |
| Factor2 | **0.49242** | **0.20410** | **0.2803** | **1.2371** |
| Factor3 | **0.28833** | **0.28556** | **0.1641** | **1.4012** |
| Factor4 | **0.00277** | **0.08887** | **0.0016** | **1.4028** |
| Factor5 | **-0.08610** | **0.07015** | **-0.0490** | **1.3538** |
| Factor6 | **-0.15625** | **0.04243** | **-0.0889** | **1.2648** |
| Factor7 | **-0.19868** | **0.06793** | **-0.1131** | **1.1518** |
| Factor8 | **-0.26662** | **.** | **-0.1518** | **1.0000** |

LR test: independent vs. saturated: chi2(**28**) = **753.29** Prob>chi2 = **0.0000**

Factor loadings (pattern matrix) and unique variances

| Variable | Factor1 | Factor2 | Factor3 | Factor4 | Uniqueness |
|---|---|---|---|---|---|
| waittime | **-0.0580** | **0.5109** | **0.0099** | **0.0043** | **0.7355** |
| constime | **0.2874** | **-0.3876** | **-0.2122** | **0.0156** | **0.7219** |
| decision_i~v | **0.4465** | **0.1496** | **-0.2604** | **0.0242** | **0.7099** |
| clarity | **0.7342** | **0.1719** | **-0.0243** | **-0.0040** | **0.4308** |
| follow_ease | **0.2028** | **-0.1551** | **0.3134** | **0.0094** | **0.8365** |
| trust_cont | **0.5747** | **-0.0461** | **0.1668** | **-0.0075** | **0.6398** |
| prov_skills | **0.6963** | **0.0103** | **0.0589** | **-0.0127** | **0.5115** |
| borrow_bin | **-0.0223** | **0.0540** | **0.2130** | **0.0400** | **0.9496** |
